# Predicting 28-Day Mortality in First-Time ICU Patients with Heart Failure and Hypertension Using LightGBM: A MIMIC-IV Study

**DOI:** 10.1101/2025.03.28.25324829

**Authors:** Junyi Fan, Shuheng Chen, Haonan Pan, Elham Pishgar, Greg Placencia, Kamiar Alaei, Maryam Pishgar

## Abstract

**Background:** Heart Failure (HF) and Hypertension (HTN) are common yet severe cardiovascular conditions, both of which significantly increase the risk of adverse outcomes. Patients with comorbid HF and HTN face an elevated risk of mortality. Despite the importance of early risk assessment, current ICU management strategies struggle to accurately predict mortality, limiting effective clinical interventions. To address this gap, we developed a machine learning model to predict 28-day mortality in ICU patients with HF and HTN.

**Methods:** We extracted data from the MIMIC-IV database, identifying a cohort of 10,010 patients with both HF and HTN, among whom the 28-day mortality rate was 9.82%. The dataset was randomly split into training (70%) and testing (30%) cohorts. Feature selection was performed using a combination of the SelectKBest and Recursive Feature Elimination (RFE) wrapper methods. To address the class imbalance, we employed the Synthetic Minority Over-sampling Technique (SMOTE). Six machine learning models were developed and evaluated: Random Forest (RF), XGBoost, LightGBM, AdaBoost, Logistic Regression (LR), and a Neural Network (NN).

**Results:** A total of 18 features were selected. The best-performing model was LightGBM, achieving an AUROC of 0.8921 (95% CI: 0.8694 - 0.9118) with a sensitivity of 0.7941 and a specificity of 0.8391 at an optimal threshold of 0.2067. External validation further demonstrated strong performance with an AUROC of 0.7404 (95% CI: 0.7130 - 0.7664).

**Conclusion:** Our proposed model achieved an AUROC improvement of 16.8% compared to the best existing study on the same topic, while reducing the number of predictive features by 18.2%. This enhanced model underscores the potential of leveraging these selected features and our LightGBM model as a valuable tool in enhancing resource allocation and providing more personalized interventions for HF patients with HTN in ICU settings.

## Introduction

Heart Failure (HF) and Hypertension (HTN) are prevalent cardiovascular conditions that pose significant public health challenges around the world. According to the American Heart Association (AHA), approximately 6.2 million adults in the United States have heart failure, with an estimated 960,000 new cases diagnosed annually^1^. Meanwhile, HTN affects nearly half of the adult population in the United States and is a major risk factor for cardiovascular morbidity and mortality^2^. Critically, patients with concurrent HF and HTN face a compounded risk of adverse outcomes, including increased hospitalizations, organ dysfunction, and mortality^3,4^. Studies have shown that patients with both conditions have a significantly higher risk of developing life-threatening complications, which often require intensive care management^5^.

In Intensive Care Unit (ICU) settings, accurately predicting the mortality risk of patients with heart failure (HF) and hypertension (HTN) is crucial to improve clinical decision making and improve patient outcomes. HF remains a significant public health concern, affecting more than 6 million adults in the United States alone, with a 5-year mortality rate exceeding 50% in severe cases^6,7^. Similarly, HTN affects approximately 45% of adults in the United States and is a major risk factor for cardiovascular events and organ failure^2,5^. Patients with comorbid HF and HTN have a particularly severe prognosis, with a substantially higher risk of adverse outcomes^3^. Despite the widespread use of risk scoring systems such as the Simplified Acute Physiology Score (SAPS) II and the Sequential Organ Failure Assessment (SOFA) score in ICU settings, these methods often demonstrate limited specificity and sensitivity, resulting in suboptimal prediction performance^8,9^. Consequently, there is a pressing need for more advanced predictive models capable of accurately assessing the mortality risk of HF patients with HTN to guide timely and personalized medical interventions.

Machine learning (ML) models have emerged as powerful tools in healthcare prediction tasks, offering improved accuracy by capturing complex, nonlinear relationships within the data. To address the limitations of traditional scoring systems, we constructed an ML-based predictive model using data from the MIMIC-IV database. Our study cohort included 10,010 patients with comorbid HF and HTN, among whom the 28-day mortality rate was 9.82%. To build a robust and interpretable model, we employed a combined feature selection strategy using SelectKBest and Recursive Feature Elimination (RFE). Given the imbalanced nature of the dataset, we applied the Synthetic Minority Oversampling Technique (SMOTE) to enhance model performance. Six ML models were explored, including Random Forest (RF), XGBoost, LightGBM, AdaBoost, Logistic Regression (LR), and a Neural Network (NN), ensuring a comprehensive evaluation.

Our results demonstrated that the LightGBM model outperformed the other ML models, achieving an AUROC of 0.8921 (95% CI: 0.8694 - 0.9118), with sensitivity and specificity of 0.7941 and 0.8391, respectively, at an optimal threshold of 0.2067. External validation further confirmed the robustness of the model, with an AUROC of 0.7404 (95% CI: 0.7130 - 0.7664). Importantly, our model achieved this strong predictive performance using only 18 selected features.

By developing a streamlined but powerful predictive model, our study advances the identification of high-risk HF patients with HTN in ICU settings. This improved risk stratification model has the potential to improve resource allocation, inform clinical decision-making, and ultimately improve patient outcomes. The alignment of our model with clinical best practices and its transparent reporting ensure its relevance for real-world applications in ICU care.

## Methods

### Data Source

The data for this study were extracted from the Medical Information Mart for Intensive Care IV (MIMIC-IV) v3.0 database^10^. MIMIC-IV is a publicly accessible critical care database that includes comprehensive identified health data from patients admitted to Beth Israel Deaconess Medical Center (BIDMC) in Boston, Massachusetts, between 2008 and 2019. The database was developed and maintained by the Laboratory for Computational Physiology at the Massachusetts Institute of Technology (MIT), in collaboration with BIDMC.

The MIMIC-IV data set used in this study is publicly available at https://physionet.org/content/mimiciv/3.0/.

### Patient Selection

The study cohort was extracted from the MIMIC-IV (v3.0) database, a publicly available critical care data set containing detailed information on admissions to the ICU of the Beth Israel Deaconess Medical Center between 2008 and 2019^11^. The database is widely used in clinical research and offers robust data on demographics, clinical measurements, and outcomes.

Our study population consisted of adult patients diagnosed with heart failure (HF) and hypertension (HTN). HF is a progressive condition characterized by the inability of the heart to effectively pump blood, leading to impaired oxygen delivery to tissues^12^. Currently, HTN is a prevalent cardiovascular risk factor and has been associated with adverse outcomes in critically ill patients^13^. Identifying patients with HF and HTN is crucial, as comorbidities have been shown to significantly increase mortality risk^14^.

To ensure a clinically relevant cohort, we applied several inclusion and exclusion criteria. First, patients with HF and HTN were identified using diagnostic codes from ICD-9 and ICD-10, which have been validated in previous cardiovascular studies as a reliable method of identifying these conditions in electronic health records.

To reduce confusion, we excluded patients diagnosed with chronic liver disease and malignancies, as these conditions are known to significantly impact mortality risk and ICU management^15,16^. Furthermore, patients under the age of 18 and over the age of 90 were excluded to ensure a more homogeneous population and to align with common practice in critical care research, where elderly patients often present with significantly different clinical profiles and treatment responses^17^.

Since our study aimed to assess the risk of mortality from the ICU, we further restricted the population to those with an ICU stay duration (LOS) between 1 and 28 days. This threshold was chosen to exclude patients who were admitted for very short periods (less than 24 hours), as these patients may not have received substantial medical intervention. To ensure data independence, only the first admission to the ICU for each patient was considered.

Following these selection steps, a total of 10010 patients were included in the final cohort for the development and analysis of the model. This process ensured that the population under investigation was representative of HF patients with HTN receiving intensive care, while minimizing confounding factors and improving the generalizability of our findings. The process is shown in Figure 1.

**Figure 1.**
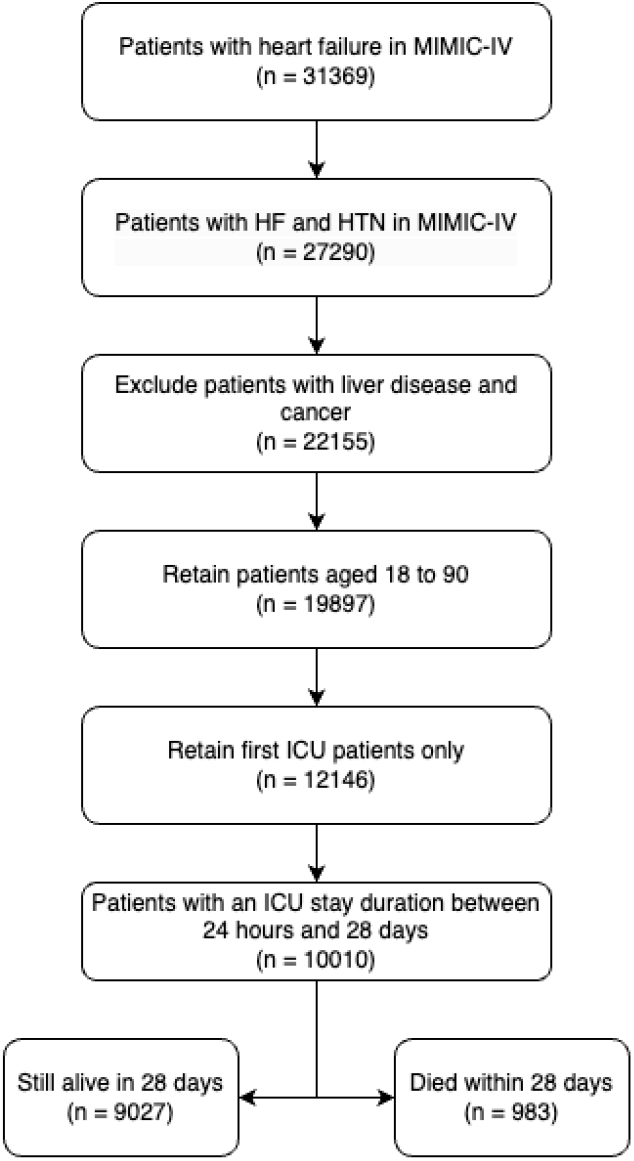
Flow chart of patient selection from MIMIC-IV dataset.

### Feature Selection Techniques

To build a robust and interpretable predictive model, we implemented a comprehensive feature selection process. Initially, more than 400 candidate characteristics were extracted from five domains: demographics, synthetic indicators, laboratory events, comorbidities, and medications. The data set was randomly divided into a training cohort (70%) and a testing cohort (30%).

Given the presence of missing values in the dataset, we adopted a K-nearest neighbor (KNN) imputation technique. Columns with more than 30% missing values were excluded. The numerical variables were imputed using KNN with *k* = 5, while the categorical variables were filled using the most frequent value, as recommended in previous studies^18,19^.

Following imputation, the numerical variables were standardized using the Z score method to ensure that they followed a normal distribution, as follows:

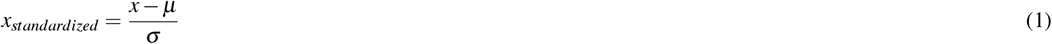

Where *x* is the original value, *µ* is the mean and *σ* is the standard deviation. Categorical variables were encoded using one-hot encoding to convert them into binary indicators for improved model performance in tree-based algorithms^20^.

Our feature selection process combined a Filter Method and a Wrapper Method to enhance model performance and minimize overfitting.

We employ the SelectKBest algorithm, which evaluates the statistical significance of each feature with respect to the target variable. The ANOVA F statistic was applied to measure the correlation between each feature and the outcome.

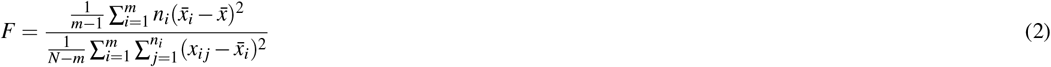

Where: - *F* is the calculated ANOVA F statistic - *m* is the number of feature groups - *n*_*i*_ is the number of samples in the *i*-th group – 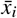 is the mean of the *i*-th group - 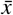 is the overall mean - *N* is the total number of samples

The top *k* features with the highest F-statistic values were selected to proceed to the next stage.

The filtered features were further refined using Recursive Feature Elimination (RFE). RFE is an iterative algorithm that repeatedly fits a model, ranks features based on their importance, and removes the least important features until the desired number of features is retained. The importance of the characteristic *x*_*i*_ was determined using the Gini Importance of the Random Forest Classifier:

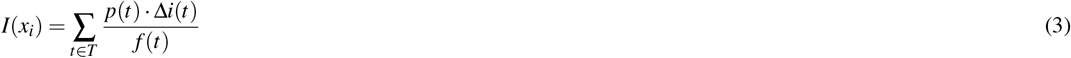

Where: - *I*(*x*_*i*_) is the importance score for the feature *x*_*i*_ - *T* is the set of all decision trees in the random forest - *p*(*t*) is the proportion of samples reaching node *t* - Δ*i*(*t*) is the Gini decrease at node *t* - *f* (*t*) is the frequency of the feature *x*_*i*_ appearing in node *t*

This combined strategy reduced the original 400 features to a streamlined set of 18 features. Our reduced feature set not only improved computational efficiency but also enhanced the model’s interpretability. In particular, this set of features achieved improved predictive performance while reducing the complexity of the model.

### Feature Details

Our study used 18 carefully selected characteristics that encompass five key domains: demographics, severity scores, laboratory markers, comorbidities, and medications. These characteristics were chosen based on their established clinical relevance and contribution to improving mortality prediction in patients with HF and HTN in ICU settings. Except for age, all features are averaged as input to the model.

Age was included as a key demographic variable due to its significant impact on clinical outcomes in critically ill patients. Studies have shown that elderly patients often face increased mortality risks in ICU environments^21^.

Two crucial clinical indicators, length of stay (LOS) and the Acute Physiology and Chronic Health Evaluation III (APS III) score, were incorporated. LOS is widely recognized as an important predictor of mortality in the ICU, where prolonged stays are associated with increased adverse outcomes^22^. APS III is a validated severity-of-illness metric derived from 17 physiological variables, each weighted by its deviation from normal values. The total score ranges from 0 to 252, with higher scores indicating increased clinical severity^23^.

Laboratory measurements played a crucial role in our feature selection process as they offer valuable insights into patients’ physiological states. Key markers included Anion Gap, Base Excess, and Bicarbonate, all of which are essential indicators of acid-base balance. An elevated atomic gap reflects metabolic acidosis, which is strongly associated with severe illness^24^. Similarly, excess base and bicarbonate imbalances are widely recognized markers of metabolic instability, which can lead to organ dysfunction and poor outcomes in critically ill patients^25^.

Base excess is calculated as:

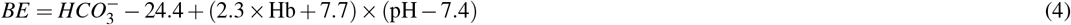

The calculation of base excess incorporates several key physiological parameters that reflect the patient’s acid-base balance. Bicarbonate concentration 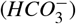 is a crucial component, representing the buffering capacity of the blood, and is measured in milliliters per liter (mEq / L). The hemoglobin concentration (Hb) is another important factor, measured in grams per deciliter (g/dL), which plays a significant role in oxygen transport and acid-base equilibrium. Furthermore, arterial blood pH (pH) is a fundamental indicator of the patient’s acid-base state, directly reflecting the concentration of hydrogen ions and the metabolic balance. These three parameters are integrated into the base excess formula to assess the deviation from normal acid-base homeostasis.

Other laboratory indicators included Bilirubin (Total), Calcium (Total), Creatinine, Glucose, Lactate, Prothrombin Time (PT), Partial Thromboplastin Time (PTT), Phosphate, Platelet Count, Urea Nitrogen, and White Blood Cells (WBC). These markers provide vital information on organ function, coagulation status, and immune response. Elevated bilirubin levels often indicate liver dysfunction and are associated with increased mortality from the ICU^26^. Creatinine levels reflect renal function, a critical factor in the prediction of mortality from the ICU^27^. Moreover, abnormalities in glucose, lactate, and phosphate have been widely studied as predictors of adverse outcomes in critically ill populations^28–30^. Meanwhile, coagulation markers such as PT and PTT are crucial to identifying clotting abnormalities, particularly in patients with severe infections or multiple organ failure^31^.

Finally, we included vasopressin administration as an essential variable to reflect hemodynamic instability. Vasopressin is frequently used to manage hypotension and has been associated with higher mortality rates when used in critical care scenarios^32^.

To evaluate multicollinearity among the selected features, we calculated the Variance Inflation Factor (VIF). VIF is a widely recognized diagnostic tool that is used to assess the extent to which the variance of a regression coefficient is inflated due to collinearity with other predictors in the model. A VIF value is computed as follows:

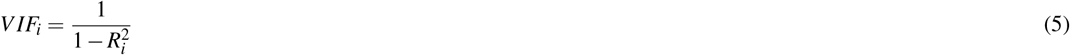

where 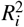 represents the coefficient of determination obtained by regressing the *i*-th predictor against all other predictors in the model 33. Generally, a VIF value that exceeds 10 indicates severe multicollinearity, while values between 5 and 10 suggest moderate multicollinearity^34^.

Our VIF analysis revealed that none of the selected features had VIF values greater than 3, indicating an acceptable level of multicollinearity across the dataset. All results are shown in Figure 2. The highest VIF values were observed for bicarbonate, excess base and creatinine, with values approaching 3. Although these values are relatively higher than other features, they are well below the common threshold of 5, suggesting that multicollinearity is not a major concern.

**Figure 2.**
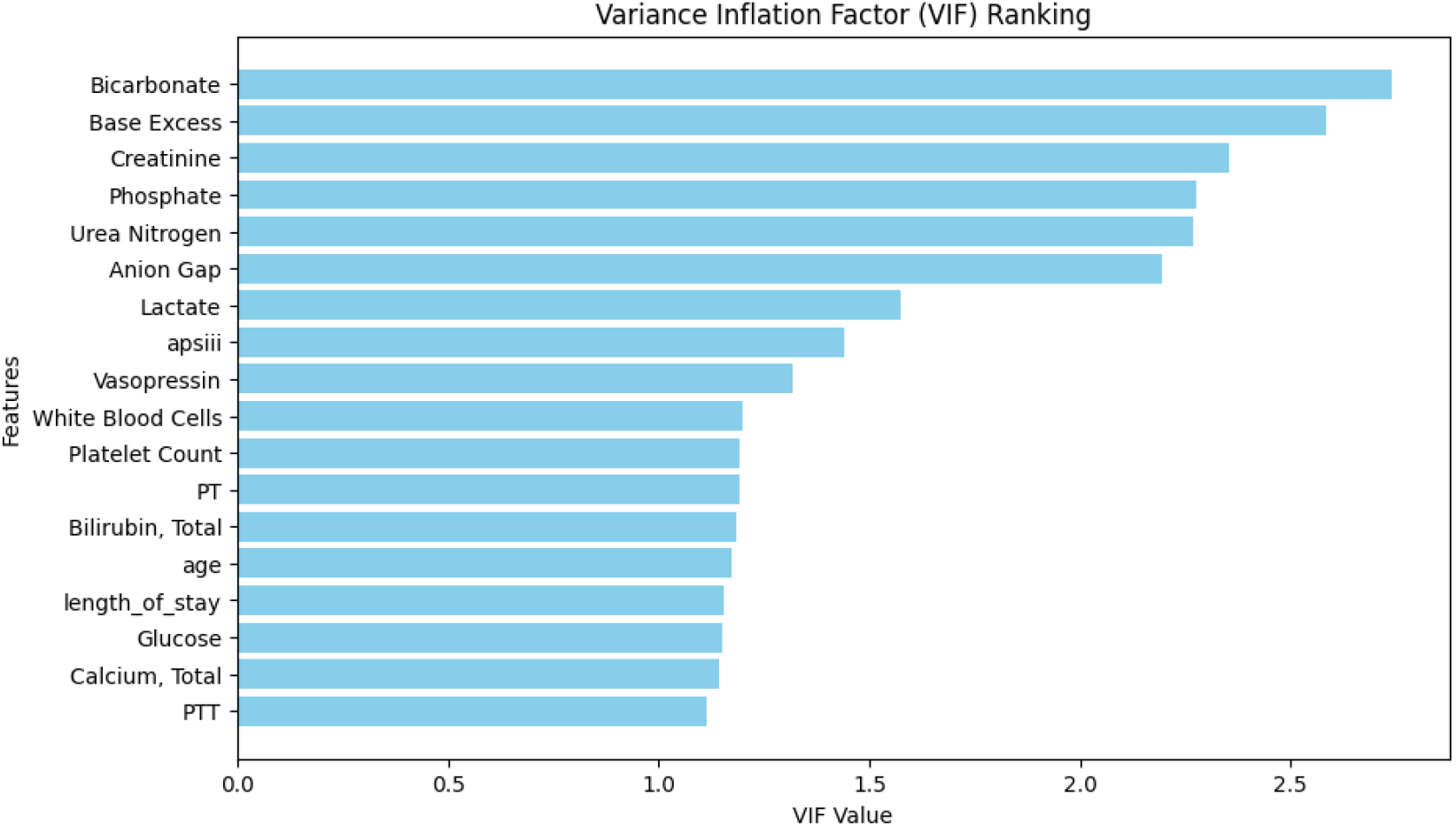
Variance inflation fact ranking results.

In contrast, features such as calcium, PTT and glucose exhibited the lowest VIF values, indicating minimal correlation with other features. Overall, the acceptable VIF values for all features ensured the stability and reliability of our predictive model.

Given that all VIF values were within acceptable limits, no feature exclusion was deemed necessary. The VIF analysis reinforced the robustness of the selected features, reducing the risk of inflated coefficients and ensuring improved model interpretability.

### Statistical Comparison

To assess the consistency of distribution between training and test sets, we performed a comparative analysis of key characteristics. Table 1 presents the mean, standard deviation, and the corresponding p-values for each feature. The p-values were calculated using independent t-tests to assess whether the mean differences were statistically significant. Most features exhibited similar distributions between training and test sets, with p values exceeding the conventional significance threshold of 0.05. This indicates a balanced distribution between the two datasets, ensuring that the training and test cohorts are comparable. However, the platelet count characteristic showed a statistically significant difference of 0.044, suggesting a potential discrepancy between the two groups. This difference may reflect biological variability or sampling effects. Despite this, the overall distribution alignment supports the validity of the model’s generalizability. The balanced distribution across most variables underscores the robustness of the data split, minimizes potential biases, and ensures that the predictive model is trained on a representative sample.

**Table 1.**
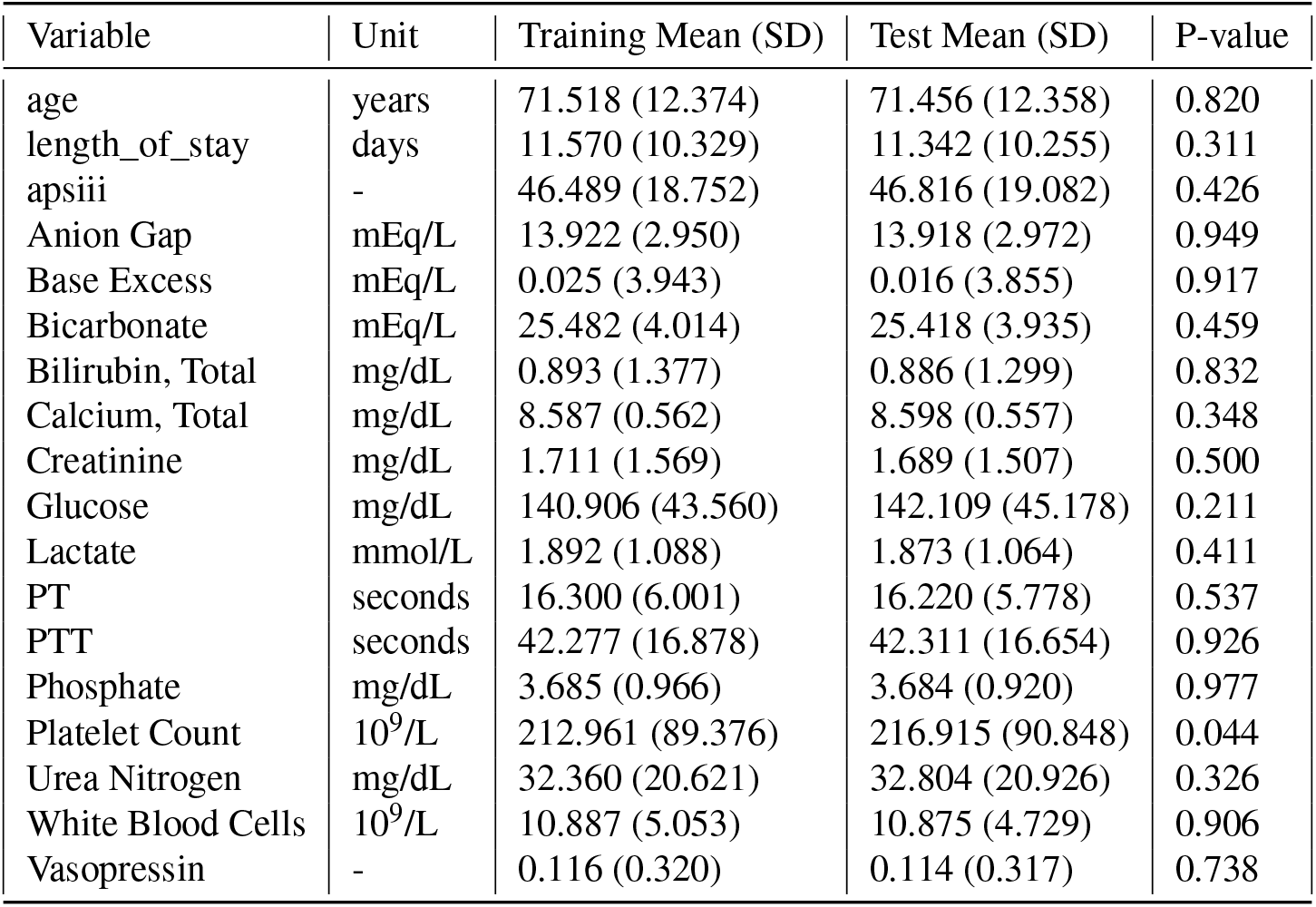
Feature Comparison Results.

| Variable | Unit | Training Mean (SD) | Test Mean (SD) | P-value |
| --- | --- | --- | --- | --- |
| age | years | 71.518 (12.374) | 71.456 (12.358) | 0.820 |
| length_of_stay | days | 11.570 (10.329) | 11.342 (10.255) | 0.311 |
| apsiii | - | 46.489 (18.752) | 46.816 (19.082) | 0.426 |
| Anion Gap | mEq/L | 13.922 (2.950) | 13.918 (2.972) | 0.949 |
| Base Excess | mEq/L | 0.025 (3.943) | 0.016 (3.855) | 0.917 |
| Bicarbonate | mEq/L | 25.482 (4.014) | 25.418 (3.935) | 0.459 |
| Bilirubin, Total | mg/dL | 0.893 (1.377) | 0.886 (1.299) | 0.832 |
| Calcium, Total | mg/dL | 8.587 (0.562) | 8.598 (0.557) | 0.348 |
| Creatinine | mg/dL | 1.711 (1.569) | 1.689 (1.507) | 0.500 |
| Glucose | mg/dL | 140.906 (43.560) | 142.109 (45.178) | 0.211 |
| Lactate | mmol/L | 1.892 (1.088) | 1.873 (1.064) | 0.411 |
| PT | seconds | 16.300 (6.001) | 16.220 (5.778) | 0.537 |
| PTT | seconds | 42.277 (16.878) | 42.311 (16.654) | 0.926 |
| Phosphate | mg/dL | 3.685 (0.966) | 3.684 (0.920) | 0.977 |
| Platelet Count | 10<sup>9</sup>/L | 212.961 (89.376) | 216.915 (90.848) | 0.044 |
| Urea Nitrogen | mg/dL | 32.360 (20.621) | 32.804 (20.926) | 0.326 |
| White Blood Cells | 10<sup>9</sup>/L | 10.887 (5.053) | 10.875 (4.729) | 0.906 |
| Vasopressin | - | 0.116 (0.320) | 0.114 (0.317) | 0.738 |

### Synthetic Minority Over-sampling Technique (SMOTE)

To address the class imbalance in our dataset, we employed the Synthetic Minority Oversampling Technique (SMOTE), a widely used method designed to enhance the representation of the minority class by synthesizing new samples based on existing data points. SMOTE achieves this by generating synthetic examples through linear interpolation between randomly selected minority class instances and their nearest neighbors^35^.

In our implementation, we first identified the minority class, defined as the class with fewer instances in the data set. Suppose *X* ∈ ℝ ^*n*×*d*^ represents the feature matrix and *Y* ∈ {0, 1} ^*n*^ is the binary target variable. Let *X*_*min*_ = {*x*_*i*_ |*Y*_*i*_ = 1}denote the set of minority class instances.

To generate synthetic samples, we employed the *k*-nearest neighbors (KNN) algorithm to identify the *k* closest points within the minority class. For each selected minority sample *x*_*i*_ ∈ *X*_*min*_, a random neighbor *x*_*neigh*_ is chosen from its nearest neighbors *k*. A new synthetic sample is then created using the following interpolation formula:

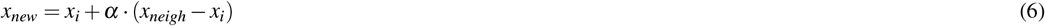

where *α* ∈ [0, 1] is a random value sampled from a uniform distribution. This ensures that the synthetic sample is located along the line segment that connects the original sample to its selected neighbor, effectively introducing variability while preserving the underlying distribution.

In our study, we set the number of neighbors at *k* = 5, a common default value that balances preservation of the local data structure and effective oversampling. Furthermore, to mitigate the risk of overfitting, the number of synthetic samples generated was restricted to 500. This controlled augmentation strategy prevents the synthetic data from disproportionately influencing the model’s decision boundary.

The SMOTE technique has been extensively validated in various medical prediction tasks and is recognized for its effectiveness in improving model performance in unbalanced datasets^36^. Using this approach, we ensured that the predictive model could better identify patterns in groups of underrepresented patients, ultimately improving the overall accuracy of the prediction.

### Model Development

In this study, we used six machine learning models to predict the risk of mortality at 28 days in ICU patients with heart failure (HF) combined with hypertension (HTN). The selected models include Random Forest, XGBoost, LightGBM, AdaBoost, Logistic Regression, and a Neural Network. Each model was tuned to achieve optimal performance, with LightGBM serving as our primary model because of its superior performance.

The Light Gradient Boosting Machine (LightGBM) is a powerful gradient boosting framework specifically designed for efficiency and scalability^37^. LightGBM leverages histogram-based learning to significantly reduce computation time and memory usage while maintaining high accuracy, making it particularly suitable for large-scale medical datasets such as MIMIC-IV. LightGBM builds decision trees in a leaf-wise manner, selecting the leaf with the greatest loss reduction at each iteration, unlike traditional level-wise growth strategies. This property enables faster convergence and improved accuracy.

The LightGBM model was configured with 500 boosting iterations to improve the model stability. The learning rate was set to 0.02 to ensure better convergence and enhance generalization. A maximum of 31 leaves was specified to allow the model to capture intricate feature interactions. The tree depth was capped at 8 to balance model flexibility and mitigate overfitting. The class imbalance was addressed using class weight balancing to give greater importance to the minority class, which was critical given the low proportion of mortality cases at 28 days in our data set. To prevent excessively small splits that might overfit noisy data, a minimum of 10 child samples was enforced in each leaf. Additionally, we employed both L1 and L2 regularization terms (reg_alpha = 0.1, reg_lambda = 0.1) to stabilize model weights and enhance feature selection. Feature sampling was applied by setting the feature fraction to 0.8, ensuring that each booster round considered only 80% of the features. Similarly, bagging techniques were applied using 80% of the data set with a bagging frequency of 5 to improve generalization performance.

Formally, LightGBM optimizes the following objective function:

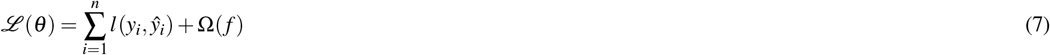

where *l*(*y*_*i*_, *ŷ*_*i*_) is the loss function that measures the difference between the predicted outcome *ŷ*_*i*_ and the true label *y*_*i*_, and Ω(*f*) is the regularization term that controls the complexity of the model to reduce overfitting.

Given the complexity of ICU data, LightGBM’s efficiency and ability to effectively model interactions among multiple clinical indicators made it the optimal choice. The model’s use of bagging, feature sampling, and class weighting provided further robustness in this high-dimensional and imbalanced dataset.

In addition to LightGBM, five reference models were implemented for comparison. Random Forest, a group learning method that aggregates multiple decision trees, was chosen for its strong performance on tabular data. XGBoost, another gradient-boosting framework, was included because of its efficiency and high performance in structured data tasks. AdaBoost was applied as an adaptive boosting model designed to combine weak classifiers into a strong model. Logistic regression, a well-established linear model, was included as a benchmark because of its simplicity and interpretability. Finally, a neural network with three hidden layers (128, 64, and 32 nodes) and a tanh activation function was used. The neural network model used the lbfgs solver for optimization, as it is effective for smaller datasets.

The LightGBM design effectively addressed the challenges associated with the prediction of mortality from the ICU in patients with HF and HTN. Its ability to handle class imbalance, nonlinear interactions, and large feature spaces aligns well with the characteristics of our dataset. This combination of capabilities makes LightGBM a strong candidate for improving risk prediction in clinical settings, particularly for patients at increased risk of adverse outcomes.

## Results

### Model Performance

The performance of the predictive models was evaluated using the Receiver Operating Characteristic Curve Area Under the Receiver Operating Characteristic Curve (AUROC), a widely accepted metric to assess the discriminative ability of binary classification models. The AUROC curve illustrates the trade-off between sensitivity (true positive rate) and specificity (false positive rate) at various threshold settings.

Our primary model, LightGBM, achieved the highest AUROC score of 0.8921, outperforming all other models included in the study. The superior performance of LightGBM can be attributed to its efficient leaf-wise growth strategy, which allows for better identification of complex patterns in high-dimensional ICU data. Furthermore, LightGBM’s ability to handle class imbalance, combined with its effective feature sampling and regularization techniques, contributed to its enhanced generalization capabilities.

Among the remaining models, XGBoost achieved an AUROC of 0.8858, closely followed by Random Forest and AdaBoost, both achieving a score of 0.8785. Logistic regression produced a slightly lower AUROC of 0.8653, while the Neural Network model had the lowest performance, with an AUROC of 0.8087. The relatively lower performance of the Neural Network may be due to the limited dataset size and the model’s increased complexity, which can result in overfitting.

Figure 3 shows the AUROC curves for all models. The LightGBM curve consistently lies above the others, demonstrating superior sensitivity and specificity over the full range of decision thresholds.

**Figure 3.**
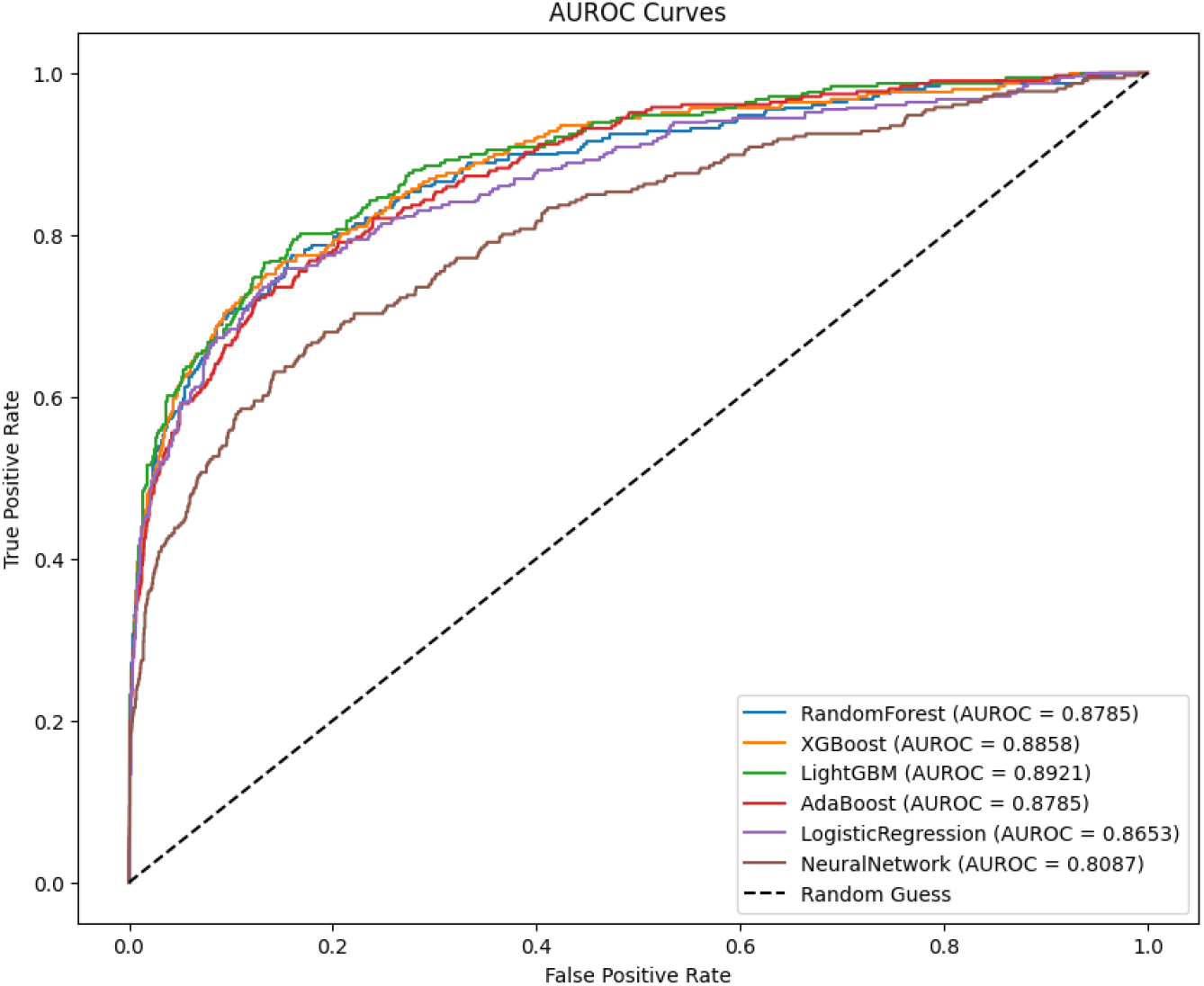
AUROC Curves for Model Performance.

Compared to a previously published model by Peng et al.^38^, which employed a neural network architecture with 22 selected features and achieved an AUROC of 0.7640, our LightGBM model not only demonstrated improved performance by 16.77% but also achieved this result with 18 selected features.

The results highlight LightGBM’s strength in clinical prediction tasks involving complex data patterns and imbalanced datasets. The performance improvement achieved with LightGBM aligns with its established reputation as an efficient and scalable gradient-boosting framework.

### Feature Analysis

We applied several ways to analyze the performance of features individually.

Ablation analysis is a technique commonly used to assess the contribution of individual characteristics to model performance by iteratively removing each characteristic and retraining the model^39^. This process helps to identify the most influential variables for prediction and improve model interpretability. The following code illustrates the ablation analysis applied in this study.

In this study, an ablation analysis was conducted on 18 selected characteristics. The results reveal that the most significant characteristics were length of stay, vasopressin, and age, causing AUROC drops of 0.0226, 0.0144, and 0.0103, respectively. This suggests that these characteristics play an important role in predicting the risk of mortality at 28 days for patients with heart failure and hypertension in ICU settings. Conversely, features like Creatinine and apsiii demonstrated minimal influence on the performance of the model, with drops in AUROC of 0.0010 and 0.0007, respectively.

These findings align with previous research that emphasizes the clinical importance of length of stay and vasopressors in mortality prediction models for critically ill patients^40,41^.

The details are listed in Table 2.

**Table 2.** Ablation Analysis Results.

| Feature Removed | AUROC Drop | Accuracy Drop | Precision Drop | Recall Drop |
| --- | --- | --- | --- | --- |
| length_of_stay | 0.0226 | 0.0123 | 0.0662 | 0.0131 |
| Vasopressin | 0.0144 | 0.0163 | 0.0846 | 0.0294 |
| age | 0.0103 | 0.0099 | 0.0532 | 0.0229 |
| Base Excess | 0.0103 | 0.0063 | 0.0351 | 0.0098 |
| Glucose | 0.0102 | 0.0070 | 0.0401 | 0.0000 |
| White Blood Cells | 0.0095 | 0.0053 | 0.0295 | 0.0098 |
| Bicarbonate | 0.0081 | 0.0050 | 0.0287 | 0.0033 |
| Phosphate | 0.0069 | 0.0067 | 0.0383 | 0.0000 |
| PT | 0.0067 | 0.0043 | 0.0255 | 0.0000 |
| Lactate | 0.0065 | 0.0037 | 0.0223 | -0.0033 |
| PTT | 0.0063 | 0.0073 | 0.0391 | 0.0196 |
| Calcium, Total | 0.0059 | 0.0017 | 0.0080 | 0.0098 |
| Urea Nitrogen | 0.0057 | 0.0027 | 0.0146 | 0.0065 |
| Platelet Count | 0.0056 | 0.0053 | 0.0326 | -0.0098 |
| Anion Gap | 0.0050 | 0.0030 | 0.0191 | -0.0065 |
| Bilirubin, Total | 0.0043 | 0.0053 | 0.0311 | 0.0000 |
| Creatinine | 0.0010 | 0.0000 | 0.0037 | -0.0163 |
| apsiii | 0.0007 | 0.0020 | 0.0114 | 0.0033 |

Permutation Feature Importance (PFI) is an effective method for assessing the contribution of each feature to the model’s predictive performance. PFI evaluates the importance of features by randomly shifting the values of a feature between observations and measuring the resulting decrease in model performance. This process is repeated multiple times to ensure stability and reduce variance. The importance of a feature *f*_*i*_ can be calculated as follows:

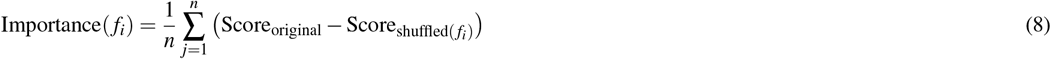

Where Score_original_ is the model performance score in the original dataset and 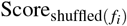 is the model performance when the feature *f*_*i*_ is randomly permuted. Higher values indicate a greater importance of the feature.

The results of the analysis of the importance of the mutation features are shown in Figure 4, where characteristics such as length of stay, age, and Vasopressin demonstrated significant importance. These features align with established clinical evidence that indicates their strong association with the risk of mortality in the ICU. The length of stay and age are widely known predictors of the mortality risk in ICU patients^42,43^, while the administration of vasopressin has been associated with severe cases requiring hemodynamic stabilization^32^. The consistency between the model’s insights and clinical findings reinforces the reliability of the LightGBM model in our study.

**Figure 4.**
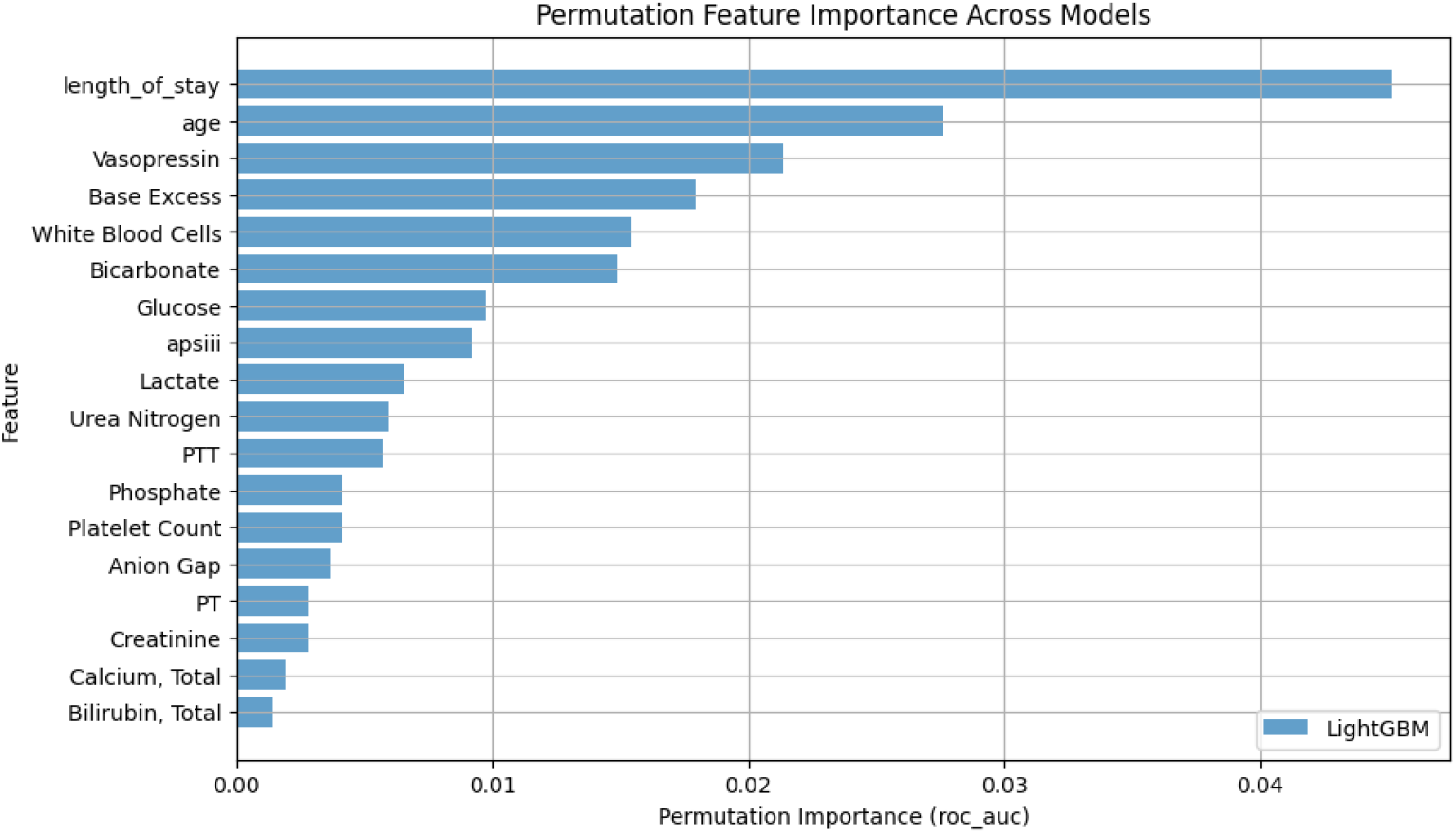
Permutation Feature Importance analysis results for LightGBM.

In both analysis methods, length of stay, age and Vasopressin were the three most important variables.

SHAP analysis (SHapley additive explanations) is a widely used method for interpreting the output of machine learning models. SHAP values are derived from cooperative game theory and assign each feature an importance value representing its contribution to the model’s prediction. For a model *f* (*x*), the SHAP value for the feature *i* in instance *x* is defined as:

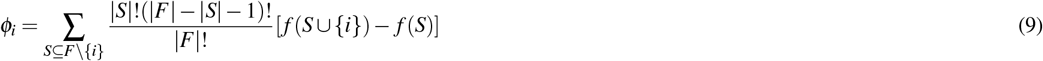

where *F* is the set of all features, and *S* is a subset of *F* without feature *i*. This formulation ensures that the contribution of each feature is fairly allocated on the basis of its marginal impact when added to a subset of features.

In our analysis, we applied SHAP to interpret the predictions of the LightGBM model. Two types of visualizations were used: the mean SHAP value plot and the SHAP summary plot.

The mean SHAP value plot, shown in Figure 5, highlights the average magnitude of SHAP values for each characteristic. This provides insights into which features have the greatest overall impact on the model’s output. As shown, *age, white blood cells*, apsiii, and length of stay emerged as the most influential features.

**Figure 5.**
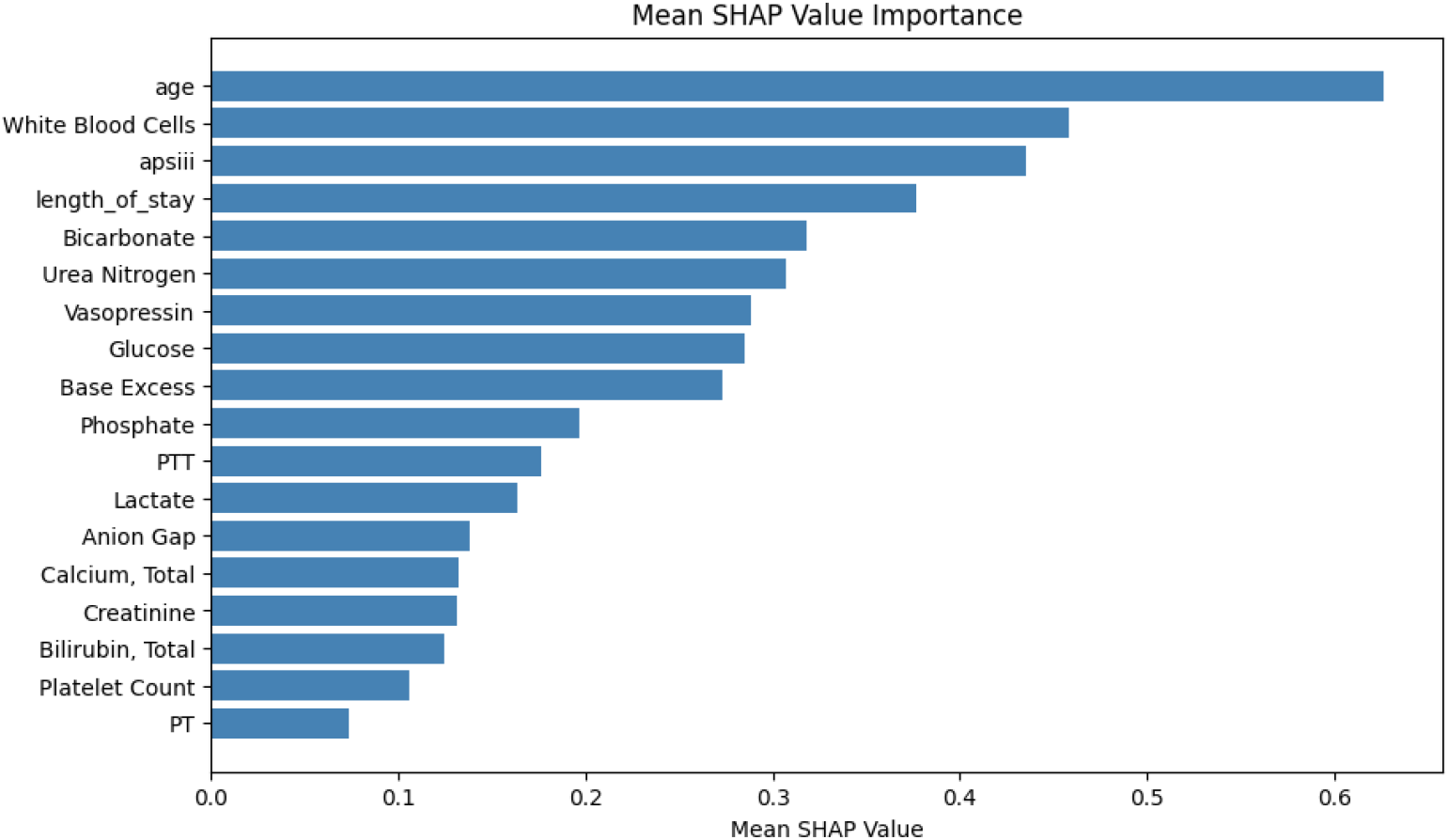
Mean SHAP Value Importance

In addition, the SHAP summary plot (Figure 6) illustrates the distribution of SHAP values for individual data points. Each point represents an individual observation, colored by the feature value (red for high values and blue for low values). The horizontal position indicates the SHAP value, demonstrating the influence of the feature on the model prediction. In particular, the length of stay exhibited strong positive and negative impacts on different observations, underscoring its complex role in predicting mortality at 28 days.

**Figure 6.**
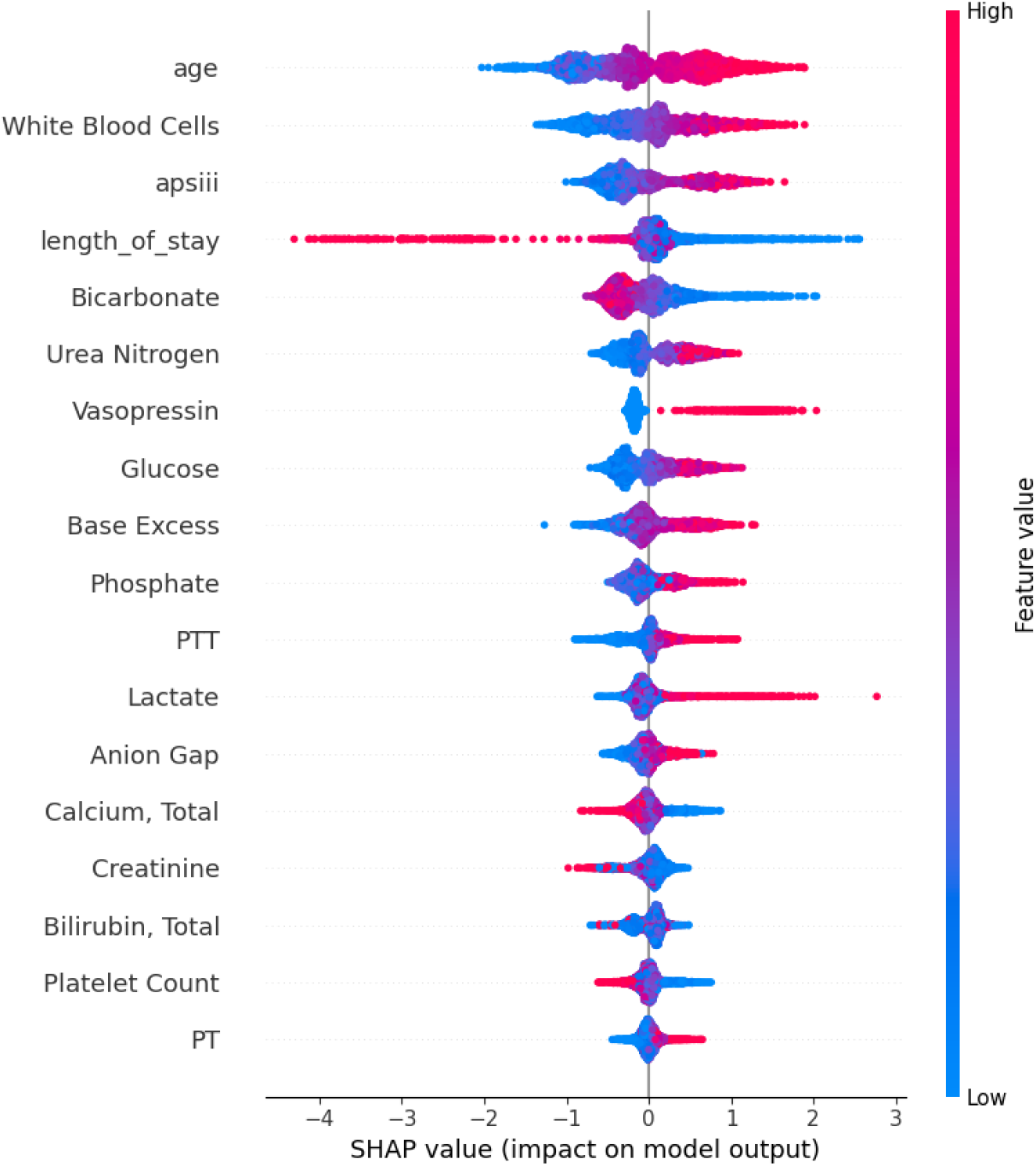
SHAP Summary Plot

The consistency between these two visualizations further validates the robustness of our selected features. Key indicators such as age, white blood cells, and length of stay consistently demonstrated high importance across the model predictions.

### External Validation

The external validation of our model was carried out using the Collaborative Research Database eICU (eICU-CRD), a large-scale critical care database that contains deidentified health data for more than 200,000 admissions to ICU throughout the United States^44^. The eICU data set was designed to support research on critical care outcomes, clinical interventions, and patient management. Its multicenter nature, which includes a variety of hospital settings and patient demographics, makes it an ideal resource to validate predictive models developed on the MIMIC-IV data set.

The eICU data set used in this study is publicly available at https://physionet.org/content/eicu-crd/2.0/.

Our model achieved an AUROC of 0.7404 (95% CI: 0.7130, 0.7664) in the eICU dataset(shown in Figure 7), with a sensitivity of 0.6302 and a specificity of 0.7195. This result closely aligns with the performance reported in the original test set of the reference study’s model, reinforcing the robustness and generalizability of our model.

**Figure 7.**
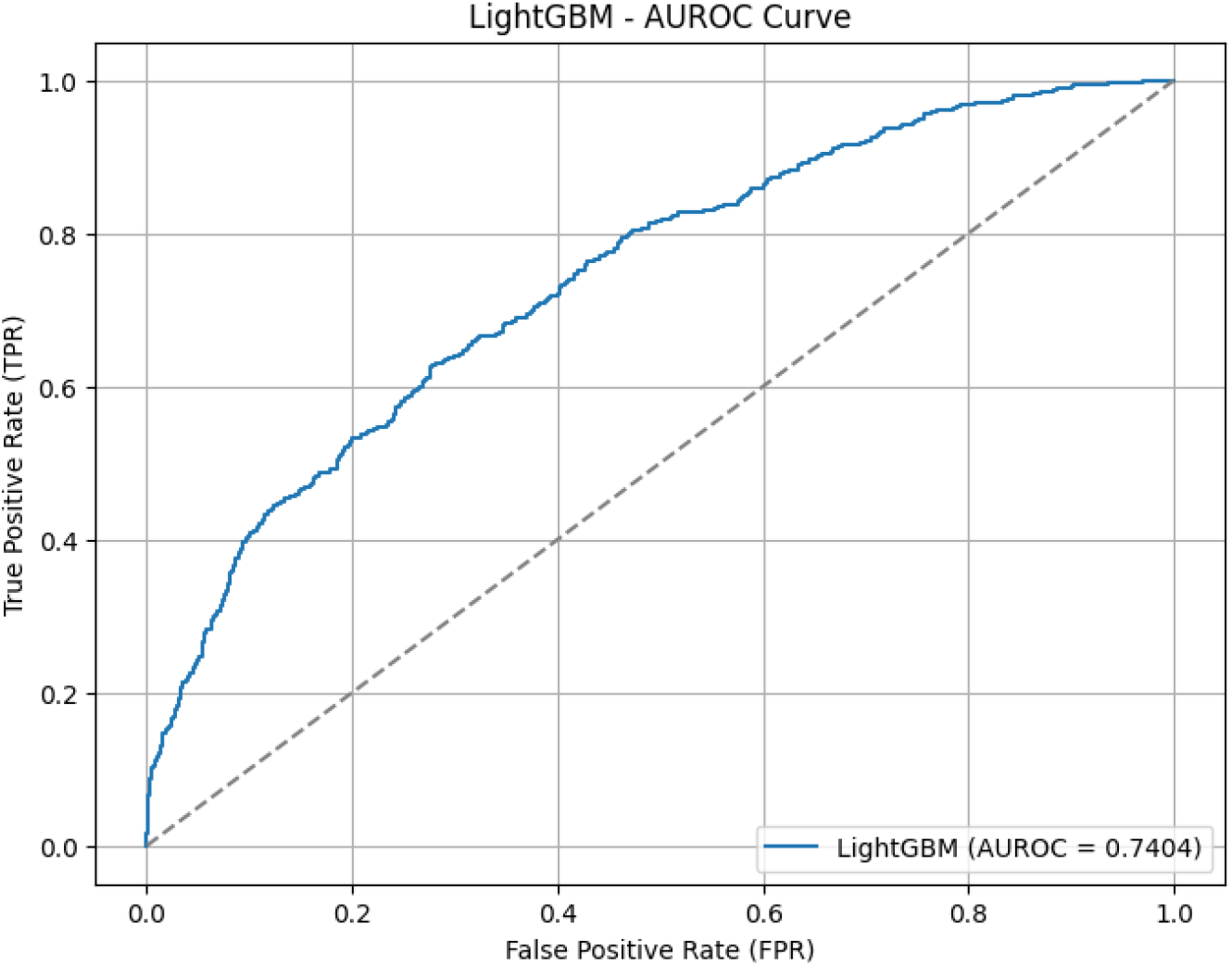
AUROC Curves for External Validation.

The consistency of our model’s performance highlights its potential utility as a reliable tool for clinical decision support in diverse healthcare settings. By validating the model on eICU data, we have demonstrated its capacity to generalize effectively, further highlighting its potential role in guiding resource allocation and risk stratification for patients with heart failure and hypertension in ICU environments.

Performance comparison of different models on internal and external datasets is listed in Table 3.

**Table 3.** Performance comparison of different models on internal and external datasets.

| Model | AUROC | 95% CI | Sensitivity | Specificity | Threshold |
| --- | --- | --- | --- | --- | --- |
| RandomForest | 0.8785 | [0.852, 0.900] | 0.7745 | 0.8409 | 0.1104 |
| XGBoost | 0.8858 | [0.862, 0.906] | 0.7516 | 0.8661 | 0.0891 |
| LightGBM | **0.8921** | **[0.871, 0.913]** | 0.7941 | 0.8391 | 0.2067 |
| AdaBoost | 0.8785 | [0.855, 0.898] | 0.7222 | 0.8747 | 0.2940 |
| Logistic Regression | 0.8653 | [0.840, 0.889] | 0.7353 | 0.8691 | 0.1248 |
| Neural Network | 0.8087 | [0.777, 0.838] | 0.6634 | 0.8254 | 0.0000 |
| LightGBM (External Validation) | 0.7404 | [0.713, 0.766] | 0.6302 | 0.7195 | 0.1976 |
| Neural Network (Peng et al.) | 0.7640 | - | 0.0280 | 0.9881 | 0.5 |

## Discussion

To predict mortality at 28 days in patients with comorbid heart failure (HF) and hypertension (HTN), we developed a machine learning model using data from the MIMIC-IV database. Our cohort included 10,010 patients with a mortality rate of 9.82%. We used a combined feature selection strategy using SelectKBest and Recursive Feature Elimination (RFE), reducing the initial feature set to 18 key indicators. To address class imbalance, we applied the Synthetic Minority Oversampling Technique (SMOTE).

Among the six models evaluated - including Random Forest, XGBoost, LightGBM, AdaBoost, Logistic Regression, and a neural network - LightGBM achieved the best performance with an AUROC of 0.8921 (95% CI: 0.8694 - 0.9118). External validation in the eICU dataset confirmed model stability with an AUROC of 0.7404 (95% CI: 0.7130 - 0.7664).

Our model demonstrated superior efficiency and performance compared to Peng et al., whose Neural Network model achieved an AUROC of 0.7640 using 22 features. Our LightGBM model improved AUROC by 16. 8% while reducing the feature count by 18.2%, underscoring its practical value in clinical decision making.

Our LightGBM-based predictive model achieved promising performance in predicting mortality at 28 days among ICU patients with heart failure (HF) and hypertension (HTN). However, certain limitations remain, both in terms of model design and clinical application.

From the perspective of model development, one limitation lies in the reliance on static baseline features extracted at ICU admission. Although these characteristics provide valuable information for early risk stratification, they do not capture dynamic changes in patient status during ICU stay, such as evolving hemodynamic patterns, electrolyte imbalances, or the impact of ongoing therapeutic interventions. Incorporating sequential or time series data could enhance the model’s adaptability to clinical progression, improving its real-time predictive value. Secondly, while our feature selection strategy effectively reduced dimensionality, some key pathophysiological markers, such as inflammatory biomarkers, myocardial injury markers, and renal function indicators, were not included in our feature set. Integrating these variables may improve the model’s ability to capture the complex interplay between multiorgan dysfunction commonly seen in ICU patients.

From a clinical application perspective, one limitation lies in the model’s reliance on retrospective data from the MIMIC-IV database. Although external validation on the eICU database confirmed model stability, both datasets are US-based ICU populations. Given regional differences in ICU protocols, treatment practices, and resource availability, further validation is necessary in international cohorts to ensure broader applicability. Furthermore, the models’ predictions may not fully account for individualized treatment decisions in the ICU setting, such as dynamic vasopressor titration, mechanical ventilation adjustments, or initiation of renal replacement therapy. These interventions, guided by clinician expertise and often tailored to each patient’s unique trajectory, introduce variability that is challenging to model using static data alone.

In the future, future research should focus on two main directions. First, improve the model design by incorporating dynamic clinical data streams, including vital sign trends, medication adjustments, and laboratory evolution, to better reflect the fast changing environment of the ICU. Second, expanding model validation across diverse geographic regions and healthcare systems is crucial to ensure the model’s generalizability and reliability. In addition, exploring clinician-in-the-loop frameworks - where the model serves as an adjunct rather than a standalone decision-making tool — may enhance its integration into real-world ICU workflows.

## Author contributions statement

Junyi Fan conceptualized the study, identified key variables, developed models, and conducted machine learning experiments. Shuheng Chen carried out the generalizability analysis. Haonan Pan performed external validation and contributed to manuscript writing. Elham Pishgar and Kamiar Alaei, as domain experts, provided medical insights regarding the topic and feature selection. Greg Placencia and Maryam Pishgar, as data science experts, advised on data sources and analytical methods. Maryam Pishgar also served as the corresponding author and supervised the overall research and manuscript development.

## Additional information

### Data Availability

The data used in this study are available from the MIMIC-IV and eICU Collaborative Research Databases, which are publicly accessible to certified researchers through PhysioNet (https://physionet.org) after completing the required data use agreements and training.

### Competing Interests

The authors declare that they have no **Competing interests** and have not received funding for this work.

## Notes

### Competing Interest Statement

The authors have declared no competing interest.

### Funding Statement

This study did not receive any funding

